# Computer-Vision Procedural Telemetry for Airway Guidance: A Public 30-Run Manikin Evidence-Package Audit

**DOI:** 10.64898/2026.06.26.26356677

**Authors:** Andrew Napier, Serge Klement, Ben Fedeles

## Abstract

**Background:** Computer vision-enabled airway workflows can turn airway video into timestamped model-observation fields, but later blinded review and training studies require source-video linkage, run identifiers, quality-control status, and app/model provenance.

**Objective:** To audit whether a public post-reconciliation 30-run manikin evidence package from a computer vision-enabled video laryngoscopy workflow preserved prespecified, video-linked procedural telemetry in structured JSON, while keeping detection accuracy, report quality, and reviewer agreement outside the current claim.

**Methods:** Thirty manikin runs were captured on a HEALTHIBLE Intubation Simulator using an IntuBlade device connected to an iPhone 15 Pro Max. Six predefined conditions were tested with five runs each in planned round-robin order by a board-certified emergency physician operator. The author-affiliated team analyzed corrected Study Metrics JSON exports, the video manifest, app/model metadata, QC fields, and the frozen package checker after reconciliation against the assigned run guide. Blinded video review, independent analysis, and report-quality adjudication were not performed.

**Results:** After reconciliation, all 30 rows contained parseable Study Metrics JSON, a companion videoFilename, run-named Drive video status, QC pass status, and corrected identifiers matching assigned row labels (30/30 for each completeness field; descriptive exact binomial 95% CI, 88.4% to 100.0%). App/model metadata were complete: appVersion 3.3.0 (75), source revision b94cd63, Navigation model, model version 31, and detection threshold 0.1. The exported JSON target-state flag was true in 25 of 25 target-condition rows (95% CI, 86.3% to 100.0%) and false in 5 of 5 no-target controls (95% CI, 47.8% to 100.0%), with zero glottic-detected frames and zero acceptable-view time in no-target controls. Among target-condition rows, median time to first model-detected glottic target was 2 seconds (IQR 1 to 3), median acceptable-view duration was 2.2 seconds (IQR 1.0 to 3.8), and median glottic visibility was 35.8% (IQR 25.8 to 45.6).

**Interpretation:** The corrected package supports a bounded formative claim: simulated airway video can be represented as specified, video-linked computer-vision procedural telemetry after documented reconciliation. It supports package completeness, traceability, and assigned-condition consistency only; it does not establish native uncorrected export reliability, computer-vision detection accuracy, report quality, reviewer agreement, training effectiveness, autonomous guidance, tube-placement confirmation, clinical efficacy, or patient outcomes.

## 1 Introduction

Endotracheal intubation is a time-sensitive procedure performed under variable anatomy, physiology, team conditions, and environmental stress. The operator must obtain a usable airway view, manage obstruction or contamination, advance the tube, confirm placement, and maintain a backup plan. Video laryngoscopy changes what the team can see. It does not remove procedural risk.

The DEVICE trial reported higher first-pass success with video laryngoscopy than direct laryngoscopy in critically ill adults [1]. That finding supports wider study of video laryngoscopy, but it leaves a separate training question unresolved. A screen can make the view visible to the team. It does not automatically create structured feedback, score procedural control, or preserve a reviewable event record.

Training has not kept pace with the video stream now available during airway management. A recorded video can help, but it remains a passive artifact if it lacks timestamps, event labels, view-state measures, and a debrief structure. The stronger opportunity is computer-vision procedural telemetry: a record of what the camera saw, what the model exported as target state, when the view was usable, when the target was lost, what confidence and visibility fields were recorded, and what human-entered checklist fields were preserved after the attempt.

This manuscript reports a 30-run manikin post-reconciliation evidence-package audit of that loop. The corrected JSON extraction verifies the chain for the reconciled 30-run package: every row links to parseable JSON, a run-named video file, build 75 app metadata, model version 31, and a unique studySessionId matching the assigned condition and replicate. The primary endpoint is corrected-package completeness and measurement readiness. Native app-export metadata defects are reported as part of the result, not hidden as incidental cleanup. The study asks whether the corrected evidence package preserves the video-linked run record closely enough to support later human review and future model development. It does not claim that guidance improves first-pass success, reduces adverse events, changes patient outcomes, or proves training effectiveness.

The computer-vision signal evaluated here is the exported run record: target-state observations, timing fields, frame counts, confidence summaries, view-state measures, and linked operator/checklist fields. It is not a clinician-adjudicated airway grade, a tube-placement decision, or a clinical success label. The system is not designed to replace airway judgment, confirm tracheal placement, diagnose airway pathology, or autonomously direct a procedure. It is a human-in-the-loop procedural guidance workflow. The contribution of this paper is the run-level computer-vision-to-JSON telemetry chain for procedural AI in airway training, with the evidence claim limited to post-reconciliation package completeness and the portions of that chain present in the corrected export package.

## 2 Related Work and Motivation

### 2.1 Video laryngoscopy and airway training

Video laryngoscopy has been studied across emergency medicine, anesthesia, critical care, and prehospital airway management. The DEVICE trial compared video and direct laryngoscopy for tracheal intubation of critically ill adults and reported higher first-pass success with video laryngoscopy [1]. That result supports the clinical relevance of video laryngoscopy, but it does not mean a screen alone creates a training system.

Airway training needs structured feedback, deliberate practice, and review. Existing scoring concepts such as percentage of glottic opening, Cormack-Lehane grade, and laryngeal-view assessment remain relevant [2,3]. Their limitation for connected video training is temporal resolution. A single post-attempt grade does not capture when the view appeared, whether it remained stable, how many corrections occurred, or whether the trainee advanced the tube during a degraded view.

Simulation studies support video laryngoscopy as a teaching surface, especially for novice operators [4-6]. Simulation reporting guidance also asks investigators to describe the simulator, scenario, participants or operators, intervention, comparator, outcomes, and data collection clearly enough for readers to judge transferability [21]. Debriefing scholarship adds a second requirement: feedback has to be anchored in observable events, not recall or global judgment alone [22]. Translational simulation further separates measurement, training, and patient-facing claims [23].

Recent airway-video studies move closer to the proposed use case. Emergency department video review has been used to measure first-pass success and timing during intubation [7]. Prehospital video laryngoscopy recordings have been used to characterize rapid sequence intubation challenges [8]. A taxonomy of hyperangulated video laryngoscopy errors shows that video can support structured performance review rather than only general debrief [9]. Neonatal work has shown that deep learning can extract procedural features, including laryngoscope insertion depth, from video laryngoscopy recordings [10]. These studies support the premise of this study: airway video can become procedural data if recording, labels, and review are designed together.

### 2.2 Procedural AI as guidance, not autonomy

Procedural AI differs from diagnostic AI. The relevant unit is not a static prediction but a sequence of actions under time pressure. A guidance overlay can change attention, timing, and operator behavior even when it does not make an explicit clinical decision. That creates a higher human-factors burden than a passive recording system.

Automation bias, over-reliance, disuse, misuse, and alert fatigue are known risks in human-machine systems [11]. In airway management, those risks are amplified because a delayed prompt, distracting overlay, or false sense of certainty could affect an attempt already under physiologic pressure. Clinical software guidance also sits near the boundary between educational support, decision support, and regulated medical-device behavior [12-14,24]. This manuscript does not claim an FDA classification, clearance, authorization, or non-device clinical-decision-support determination.

Reporting guidance for clinical AI makes the same point from a study-design perspective. CONSORT-AI and SPIRIT-AI call for clear descriptions of inputs, outputs, integration points, failure handling, and human interaction [15,16]. DECIDE-AI focuses on early-stage clinical evaluation, where the question is often whether a system can be used safely and understood correctly before an outcomes trial is justified [17]. Airway-management guidance emphasizes oxygenation, capnography, attempt limitation, escalation planning, and failed-airway rescue as clinical safety requirements [25,26]. A procedural airway overlay belongs in an early evaluation category until it has been tested against video review, human-factors outcomes, and predefined safety thresholds.

## 3 Computer Vision Telemetry Workflow and Evidence Boundary

The workflow description is limited to the system elements needed to reproduce and audit the 30-run corrected package. The workflow has five operational layers and one output layer: video input, computer vision observations, guidance state, event telemetry, exported operator/checklist fields, and debrief or validation output. The implementation context is an iOS connected airway guidance workflow developed by the study team [18]. The reproducibility record for this analysis is app build 3.3.0 (75), source revision b94cd63, Navigation model, model version 31, detection threshold 0.1, and the archived telemetry schema. The private implementation repository is cited only to define the source boundary [18]. The implementation is a testbed for the evidence-chain study. It is not evidence that any model is clinically effective, and this report does not disclose or validate the model architecture, weights, or training data.

A package telemetry transparency card is included in the public evidence package. It names the intended use as simulator training-feedback telemetry, lists the observed input source and exported output fields, records the threshold and model identifiers available in the package, and states what is not disclosed or not inferred. That card is a transparency artifact for this package, not a clinical model card or a substitute for a future model card with architecture, training data, validation labels, latency testing, drift monitoring, update policy, and intended-use risk analysis.

Figure 1 shows a representative four-panel simulator-airway sequence from the controlled source videos. It is included to show the visual input, overlay prompts, tube passage, and app-view context used in the workflow, not as an analyzed outcome. The evidence-chain schematic in Figure 2 summarizes how those video observations are separated from human-entered fields, derived outputs, and clinical-claim boundaries.

**Figure 1.**
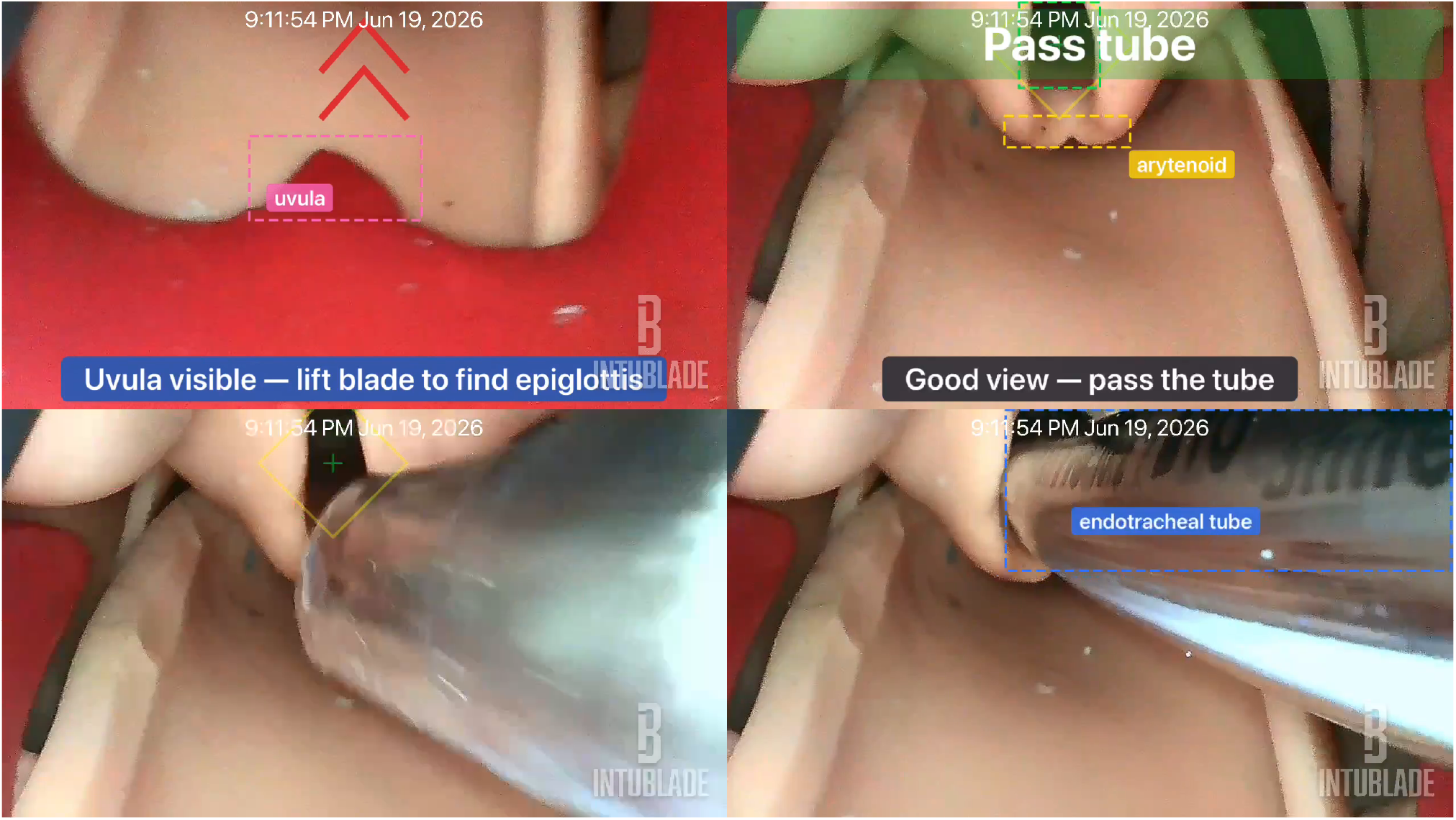
Representative four-panel simulator-airway guidance sequence from run C5-R5 showing uvula-directed feedback, target/arytenoid guidance, tube advancement, and endotracheal-tube overlay in the captured app view. Branding and timestamp context are retained. This is visual context only, not detection-accuracy, tube-placement, training-effect, clinical-efficacy, or measurement-validity evidence.

**Figure 2.**
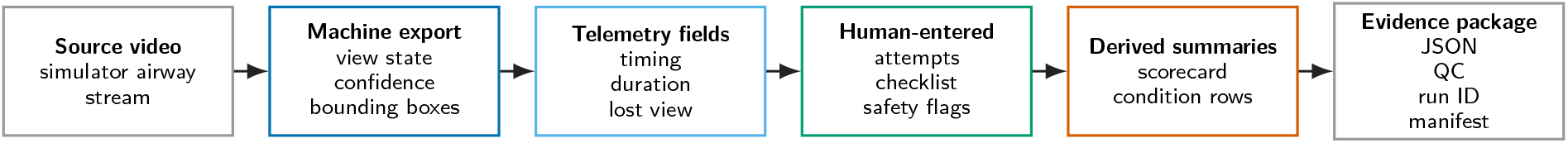
Computer-vision telemetry evidence chain. The schematic shows how a simulator airway video is represented as machine-exported observations, event telemetry, human-entered fields, derived summaries, and an auditable run package. This audit checks traceable telemetry, not detection accuracy, tube-placement confirmation, training effect, clinical performance, or patient outcomes.

The evidence unit is a run package, not a product demonstration or an aggregate score. A complete future measurement-validity package would link source video, computer vision observations, guidance-state telemetry, exported operator/checklist fields, generated report, JSON export, app version, source revision, active model type, active model version, blinded labels, adjudication, and failure notes. The observed JSON extraction verifies the JSON, video, app/model, and corrected run-ID portions of that chain. Blinded labels, adjudication, and report-quality files are not reported from the JSON-only package. The study therefore evaluates corrected-package telemetry traceability, not independent computer-vision detection performance.

The input layer captures the airway video stream used for the run. In simulator mode, a video file can be replayed through the same detection and reporting path. This allows repeatable testing before cadaver or clinical settings are considered. The computer vision layer detects airway-view states used for training feedback. It is not complete airway-scene understanding, tube-placement confirmation, or autonomous airway assessment. The primary machine outputs are glottic-opening or target-state observations with timing, confidence, frame-count, and bounding-box metadata. Safety-relevant events such as tube placement, capnography confirmation, escalation, and esophageal concern remain outside machine inference and are represented only by exported operator/checklist fields when entered.

The guidance layer renders a low-profile marker around the detected glottic opening and records view-centeredness states. The operator keeps control of the procedure. The overlay does not insert the tube, confirm the airway, decide when to abandon an attempt, or judge patient safety. During simulation it can show target presence, target centeredness, directional correction state, target loss, and confidence behavior. It cannot show tube placement, physiologic status, capnography, escalation decision, or clinical success. False target, missed target, delayed overlay, distracting overlay, over-reliance, disuse, and alert fatigue remain human-factors hazards for later testing; this package tests exported evidence-chain integrity rather than deployment readiness.

The telemetry layer records session start and end, first model-detected glottic-target timestamp, acceptable-view duration, longest centered streak, guidance-correction count, lost-view count, processed frame count, glottic-detected frame count, average detection confidence, maximum glottic width ratio, screen-view POGO proxy, screen-view Cormack-Lehane proxy, time to best view, attempt count, navigation events, active model type, active model version, and exported operator/checklist fields. Attempt count is nullable by design because this model does not infer tube entry and withdrawal; nil means not recorded, not one attempt.

The reporting layer converts telemetry into a session report, Study Metrics JSON export, and trainer-facing debrief summary. The report view includes session summary, study export, key metrics, view and technique assessment, trainer checklist, and coaching summary. The technique assessment is rule-based. It uses explicit threshold rules over session metrics rather than a generative language model.

The validation boundary has six requirements before any cadaver or clinical extension: human control, suppressibility, conservative scope, mode clarity, traceability, and evidence gating. The current package documents only the traceability and corrected-package readiness portion of that boundary. Later studies require a named gov-ernance workflow covering access control, deidentification, retention limits, local review, operator consent where applicable, trainer oversight, suppressibility, and a written boundary that the system does not confirm tube placement or direct patient care. A governance-readiness note in the evidence package records current status, required artifacts, owners, triggers, stop conditions, and controlled-access video request fields before any cadaver, clinical quality-improvement, or patient-facing extension.

## 4 Methods

This formative post-reconciliation evidence-package audit used a 30-run manikin dataset to test whether a computer vision-enabled video laryngoscopy workflow could yield a complete, video-linked Study Metrics JSON evidence package suitable for later blinded video review. The unit of analysis was the run. The primary end-point was corrected-package completeness, defined as parseable JSON, a companion videoFilename, run-named Drive video presence, app/model metadata, QC status, and alignment between corrected JSON run identifiers and the prospectively assigned Sheet row labels. Native app-export metadata integrity was analyzed as a failure mode, and corrected-package scoring began after documented reconciliation against the prospectively assigned run guide. The validation build, model configuration, telemetry schema, and study export were locked before JSON scoring. For binary package checks, descriptive exact binomial 95% confidence intervals are reported to show precision only; they are not inferential tests of clinical performance. The package checker verifies the frozen corrected package only; raw app-export metadata defects are reported separately in the QC Summary and are not converted into evidence of native export reliability. The blinded labeling guide and adjudication rules define the next review step, but the reviewer-label fields were not populated in the JSON-only source analyzed here. The observed results reported here come from the corrected export package; reviewer-labeled agreement and report-quality scoring are not reported.

The manuscript is reported as an early-stage procedural AI and formative simulation study. DECIDE-AI informs reporting of inputs, outputs, workflow insertion points, human interaction, failure handling, and safety boundaries [17]. Simulation reporting guidance informs reporting of simulator context, scenario conditions, operators, outcomes, and data collection [21]. CONSORT-AI and SPIRIT-AI are adjacent AI-reporting references because this study is not a randomized clinical trial or clinical trial protocol [15,16].

### 4.1 Simulation setting and run conditions

The validation dataset contained 30 manikin intubation videos acquired on a HEALTHIBLE Intubation Simulator Endotracheal Simulator (Amazon ASIN B0BMLPRPYY) using an IntuBlade device connected to an iPhone 15 Pro Max: six predefined conditions with five runs per condition. Runs were captured in planned round-robin order from C1-R1 through C6-R5 rather than randomized order. A board-certified emergency physician with more than 20 years of intubation experience followed the same condition guide used to build the capture sheet. Capture, data extraction, metadata reconciliation, figure generation, and manuscript analysis were performed by the study team; no independent operator, external analyst, or blinded adjudicator contributed to the observed JSON-only results reported here.

C1 was a clean-lens, normal approach run with early glottic acquisition and stable view. C2 intentionally held the glottis off-center for 2 to 3 seconds before correcting and finishing. C3 acquired the glottis, deliberately lost the view for 1 to 2 seconds, then reacquired and finished. C4 introduced rapid motion to simulate chest-compression-like visual motion. C5 made tube passage past the vocal cords clearly visible and kept recording through completion. C6 was a no-target control: no glottic opening visible, no tube passage, and 15 to 30 seconds of mouth, tongue, palate, black-field, or other non-target video.

Runs were excluded only if the source video was missing, the run ID could not be reconciled with the export, or the file was corrupted before analysis. Excluded runs were to be listed before replacement or resampling. No run in the corrected 30-row export package was excluded: all 30 rows had parseable JSON, companion videoFilename, run-named Drive video presence, and QC status OK.

### 4.2 Source package and reproducibility

For this validation dataset, the reproducibility record was app build 3.3.0 (75), source revision b94cd63, Navigation model, model version 31, and detection threshold 0.1. Each run package included the Study Metrics JSON export, companion video filename, Drive video-upload status, assigned condition, replicate number, app/model metadata, timing and visibility summaries, and exported operator/checklist fields. The public package documents model identity and threshold as operational metadata, but it does not provide model architecture, training data, weights, or frame-level ground-truth labels.

The archived evidence package preserves the corrected Sheet export, corrected raw JSON exports, run manifest, QC Summary, correction ledger, package hash manifest, data dictionary, provenance note, observed-results summary, package telemetry transparency card, governance-readiness note, app build, source revision, model version, analysis script, and package-check script. The package checker passed after data lock. Public reproducibility is therefore at the corrected JSON, manifest, QC, correction-ledger, package-hash, package-check, and figure-generation level. The public repository does not support independent reproduction of video-to-JSON extraction, model inference, or source-video content verification because the implementation repository, model internals, source videos, and report screenshots remain controlled-access or private artifacts. Video evidence is referenced by filename in the manifest, and no manuscript endpoint depends on report screenshots. Reviewer labels are reserved for controlled-access blinded-review artifacts after that work is performed.

### 4.3 Outcomes and analysis

The observed JSON-extraction endpoints were parseable Study Metrics JSON export, companion videoFilename presence, Drive video-upload status, app and model metadata completeness, run-ID traceability, assigned-condition consistency of exported target-state fields, exported esophageal-related checklist fields, time to first model-detected glottic target, acceptable-view duration, glottic visibility, average confidence, screen-view POGO proxy, and exploratory composite score. Export completeness used 30 eligible rows as the denominator. A field was considered complete when it was present and parseable in the corrected export or manifest. Run-ID matching required corrected studySessionId, condition, and runNumber to match the assigned row label. Expected target/no-target behavior meant JSON target-state flag=true for predefined target-condition rows and JSON target-state flag=false with zero glottic-detected frames and zero acceptable-view duration for predefined no-target controls. This assigned-condition check is not a sensitivity, specificity, or frame-level false-positive/false-negative estimate because the source videos were not independently adjudicated in this analysis.

JSON-derived timing and view fields were treated as machine-exported measurements, not adjudicated ground truth. Time to first model-detected glottic target was seconds from session start to first glottic-opening detection. Acceptable-view duration was cumulative model-defined acceptable-view time in seconds. Glottic visibility was glottic-detected frames divided by total processed frames. Average confidence was the mean confidence across glottic detections when available. Screen-view POGO proxy, screen-view Cormack-Lehane proxy, and composite score were app-generated descriptive fields and were not treated as validated clinical measures.

Reviewer-dependent endpoints remain prespecified but are not scored from the Sheet alone. These include condition-label agreement against blinded labels, time-to-first-glottic-view error against video timestamp labels, acceptable-view timing error, correction-count agreement, lost-view-count agreement, tube-passage event agreement, no-target false-target review on source video, and report-quality score.

Export completeness, videoFilename presence, video-upload status, app/model metadata completeness, run-ID matching, unique studySessionId values, no-target behavior, and exported esophageal-related checklist fields are summarized as counts out of eligible rows. Continuous JSON-derived fields are summarized with median and interquartile range using the same linear percentile convention across manuscript tables and package summaries. No inferential tests were planned or performed. Reviewer-dependent agreement statistics are intentionally not calculated from the Sheet alone. C6 No-target runs are analyzed separately because first model-detected glottic target and acceptable-view timing are not defined when no glottic target is present.

Human-factors outcomes belong in a later operator study because repeated attempts by the same participant are not independent. The 30-run sample size is appropriate for formative validation and failure-mode discovery. It is not powered for clinical efficacy. Five rows per intended condition can expose export, traceability, no-target, and scorecard failures before larger cadaver or operator studies.

Substantive empirical claims in the Results and Discussion are anchored by bracketed citations, numbered equations, or the D/E/A labels in Table 1. The labels are audit anchors: D-labels refer to supplied data or package files, E-labels to analysis rules, and A-labels to explicit study-boundary assumptions. Narrative background claims remain citation-traced and are not treated as observed results.

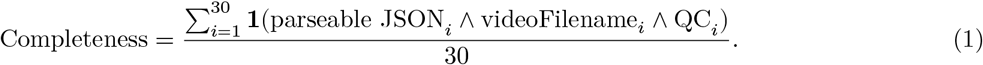

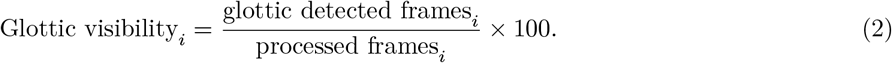

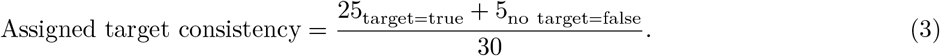

**Table 1.** Traceability key for claims.

| Label | Trace source |
| --- | --- |
| D1 | Apparatus: HEALTHIBLE manikin; IntuBlade device; iPhone 15 Pro Max. |
| D2 | Run guide: C1–C6; five runs per condition. |
| D3 | Acquisition: planned round-robin order; single board-certified emergency physician. |
| D4 | Package files: corrected JSON; Drive manifest; QC Summary; checker. |
| D5 | Corrections: 7 studySessionId; 7 runNumber; 2 condition; build-74 metadata. |
| D6 | Completeness: 30 JSON; 30 videoFilename; 30 videos; 30 QC pass. |
| D7 | App/model: 3.3.0 (75); b94cd63; Navigation; model 31; threshold 0.1. |
| D8 | Target/no-target: 25/25 true; 5/5 false; C6 zero frames/time. |
| E1 | Summaries: CSV medians/IQR by linear percentile convention. |
| E2 | Denominator: 30 eligible corrected rows. |
| E3 | Scope: assigned-condition check, not accuracy. |
| A1 | Not performed: blinded video review, independent analysis, report adjudication. |
| A2 | Access boundary: source media controlled-access; implementation private. |

## 5 Results

The Sheet contained 30 corrected Study Metrics JSON exports. The correction process was part of the endpoint: original app exports had metadata defects, and corrected-package scoring was performed only after reconciliation against the prospectively assigned run guide. Native app-export metadata integrity therefore failed for build metadata in 30/30 rows and for run identifiers in a subset of rows;^D5^ the endpoint reported here is frozen corrected-package completeness, not native export reliability. After correction, all 30 exports parsed, included a videoFilename, and had an uploaded run-named video in the Drive capture folder. The exports were aligned for run metadata: 30/30 studySessionId values matched the visible Run ID, 30/30 condition fields matched the assigned C value, 30/30 runNumber fields matched the assigned R value, and all 30 studySessionIds were unique.^D6E2^ All rows used appVersion 3.3.0 (75), source revision b94cd63, Navigation model, model version 31, and detection threshold 0.1.^D7^ Reviewer agreement, video-timestamp error, event-count agreement against blinded labels, and report-quality scores are not reported because those reviewer/adjudication fields were not present in the Sheet.^A1^

### 5.1 JSON completeness, video linkage, and traceability

The primary export path was complete in the corrected Sheet: 30/30 rows contained parseable Study Metrics JSON, 30/30 JSON exports included a videoFilename, 30/30 run-named videos were present in Drive, and 30/30 rows passed QC (descriptive exact binomial 95% CI, 88.4% to 100.0%).^D6E1^ Run-package traceability was complete after correcting app-export metadata against the assigned row labels: 30/30 studySessionId values matched the visible Run ID, 30/30 condition fields matched C, 30/30 runNumber fields matched R, and all 30 studySessionIds were unique.^D6^ The live Sheet and frozen evidence package preserve the correction log in the QC Summary tab and provenance note.

Figure 3 shows all supplied run-level stress values and Table 2 summarizes the same condition-level medians. The target-state flag column in Table 2 reports assigned-condition consistency only and is not a sensitivity, specificity, or frame-level false-positive/false-negative estimate.

**Table 2.** Condition-level JSON-derived results.

| Condition | Runs / JSON target-state flag | Median first target / acceptable view | Median visibility / lost views | Median composite score |
| --- | --- | --- | --- | --- |
| C1 Clean | 5; 5/5 true | 3.0 s / 3.1 s | 43.2% / 3.0 | 82 |
| C2 Off-center | 5; 5/5 true | 3.0 s / 6.0 s | 48.1% / 8.0 | 84 |
| C3 Lost/reacq. | 5; 5/5 true | 3.0 s / 2.2 s | 26.6% / 4.0 | 84 |
| C4 Degraded | 5; 5/5 true | 1.0 s / 0.8 s | 25.4% / 16.0 | 82 |
| C5 Tube passage | 5; 5/5 true | 1.0 s / 1.5 s | 45.5% / 3.0 | 84 |
| C6 No-target | 5; 5/5 false | not applicable / 0.0 s | 0.0% / 0.0 | 10 |

**Figure 3.**
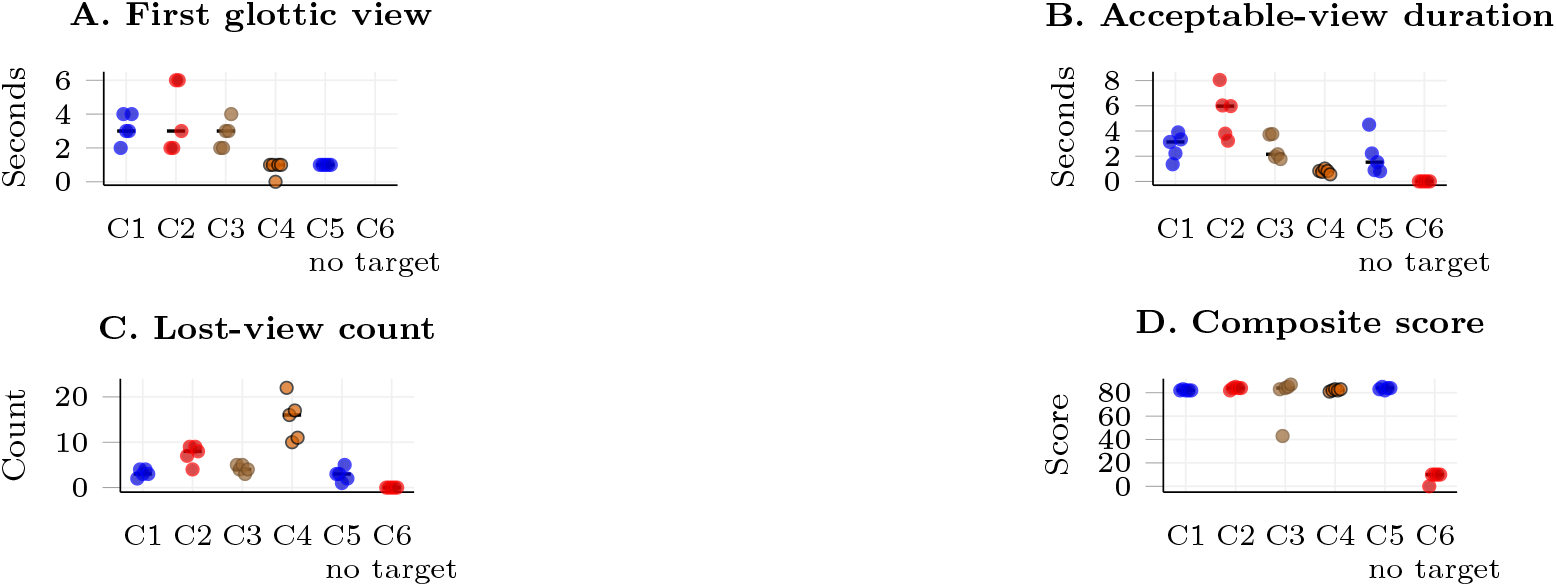
Run-level stress signatures by predefined condition. Points show individual observed runs; black ticks show medians. C6 is the no-target control; panel A has no C6 points because first glottic view is not defined when no glottic target is present.

### 5.2 Target/no-target behavior, esophageal-related fields, and exploratory scores

The JSON target-state flag was true in 25 of 25 target-condition rows (descriptive exact binomial 95% CI, 86.3% to 100.0%) and false in 5 of 5 no-target controls (descriptive exact binomial 95% CI, 47.8% to 100.0%), with zero glottic-detected frames and zero acceptable-view time in the no-target controls.^D8E3^ None of the 30 JSON exports had a positive esophageal-related checklist field: esophagealIntubationDetected and oesophagealConcern were false, and esophagealInsertionCount was zero. These findings describe exported-field behavior under predefined manikin conditions, not adjudicated detection accuracy, sensitivity, specificity, frame-level false-positive or false-negative rates, tube-placement status, or patient-safety performance.^A1^ Among target-condition rows, median time to first model-detected glottic target was 2 seconds (IQR 1 to 3), median acceptable-view duration was 2.2 seconds (IQR 1.0 to 3.8), median glottic visibility was 35.8% (IQR 25.8 to 45.6), median average confidence was 0.61 (IQR 0.56 to 0.68), and median exploratory composite score was 83 (IQR 82 to 84).^E1^ The composite score and screen-view POGO proxy remain descriptive training-feedback fields.

Figure 4 displays the target/no-target separation used for the assigned-condition consistency statement in Equation 3.

**Figure 4.**
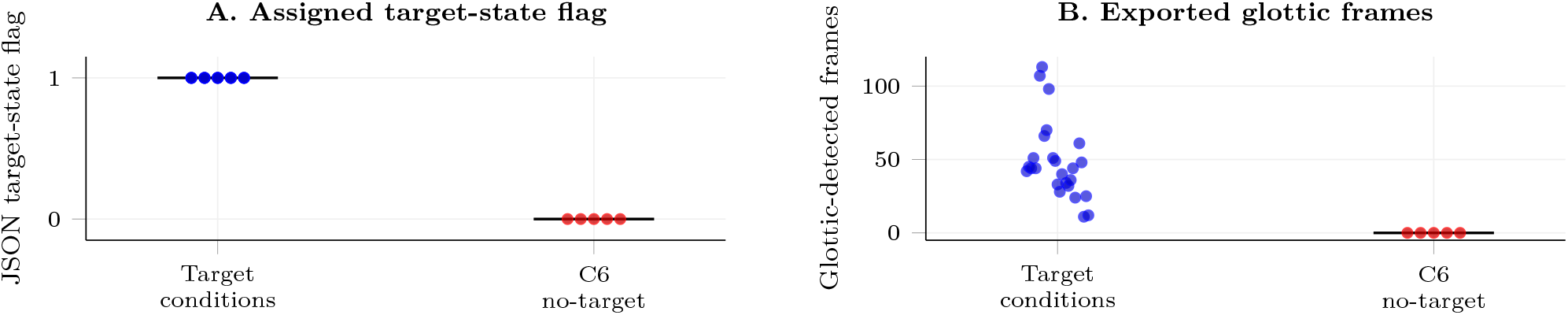
Target/no-target JSON behavior and interpretation limits. Target-condition rows had target-state true (25/25); C6 controls had target-state false (5/5) and zero glottic-detected frames. This is assigned-condition consistency only, not detection accuracy or clinical performance.

### 5.3 Data reconciliation

The QC Summary records the correction step. Original app exports had seven studySessionId mismatches, seven runNumber mismatches, two condition mismatches, and all rows initially reported build 74. Those raw JSON metadata fields were corrected by deterministic lookup against the prospectively assigned Sheet row labels, which served as the prespecified run log for condition and run number. Row placement is author-attested in the current package rather than independently adjudicated. No JSON-derived timing, frame-count, confidence, visibility, target-state, score, or checklist field was edited for the manuscript analysis. These defects mean the study should not be read as native, uncorrected end-to-end export-reliability validation. Rather, it evaluates whether the corrected evidence package preserves a complete and auditable computer-vision telemetry record after documented reconciliation. The repository preserves the corrected raw JSON exports, corrected Sheet export, QC Summary with old metadata values, provenance note, and structured correction ledger; the correction history is therefore auditable but not independently adjudicated in this report. After correction, all 30 rows had unique studySessionId values; all 30 matched the visible Run ID, assigned C value, assigned R value, and appVersion 3.3.0 (75). No-target detection failure was not observed in the JSON exports. All five C6 rows had zero glottic-detected frames. Reviewer agreement and report quality are not reported from the JSON-only Sheet.

## 6 Discussion and Limitations

The corrected 30-run package supports a formative telemetry finding: airway video from predefined manikin conditions can be represented as complete, source-linked computer-vision procedural telemetry after documented metadata reconciliation. Every corrected row had parseable Study Metrics JSON, every JSON export included a videoFilename, every run-named video was present in the Drive manifest, app and model metadata were complete, and every corrected studySessionId matched the assigned run label.^D6–7E2^ The result is narrow by design. It shows that the attempt can be preserved as a reviewable data object rather than reduced to a small number of manually entered fields. It does not show that the native uncorrected export was error-free, and it does not reproduce the private video-to-JSON extraction path.^A1–2^

The clinical context matters because video laryngoscopy already changes what the operator and instructor can see. The DEVICE trial showed better first-pass success with video laryngoscopy than direct laryngoscopy in critically ill adults [1], and prior simulation studies have used video review to improve novice airway performance [4-6]. This manuscript does not test those clinical or training outcomes. It addresses the step before them: whether a connected video laryngoscopy workflow can produce a complete run-level evidence package suitable for blinded review, debrief, and later model development.

The target/no-target pattern is the cleanest package-level consistency check in the observed dataset. The JSON target-state flag was true in 25 of 25 target-condition rows (descriptive exact binomial 95% CI, 86.3% to 100.0%) and false in 5 of 5 C6 no-target controls (descriptive exact binomial 95% CI, 47.8% to 100.0%) with zero glottic-detected frames and zero acceptable-view time.^D8E3^ That separation shows that exported target-state fields were internally consistent with predefined target/no-target conditions under this manikin protocol. It should not be read as detection accuracy because the source videos were not independently adjudicated in this analysis. It does not prove independently adjudicated target discrimination, tube-placement detection, prevention of esophageal intubation, or clinical guidance.^A1^

The condition-level stress signal is also useful for debrief design. C4 degraded-view runs carried the highest lost-view burden, while C1 clean view, C2 off-center correction, C3 lost-view/reacquisition, C5 tube passage, and C6 no-target controls retained distinct descriptive profiles (Figure 3; Table 2).^D4E1^ These differences move the record beyond a binary pass/fail label. Time to first model-detected glottic target, acceptable-view duration, lost-view count, centered-view streaks, confidence, visibility, and screen-view POGO proxy are the computer-vision telemetry artifact produced by the workflow. They describe how the exported view state changed during the run. They remain descriptive unless blinded reviewers and prospective studies validate them as clinical or training measures.^A1^

Source labeling is the main safety control. Some fields are machine observations, some are exported human-entered or checklist fields, and some are derived debrief outputs. Attempts, capnography, tube confirmation, esophageal concern, and escalation remain human-entered checklist inputs rather than machine inferences. Screen-view POGO proxy and screen-view Cormack-Lehane proxy are computer-vision approximations, not clinician-confirmed grades or difficult-airway predictors. The composite score is a candidate debrief summary, not a patient-safety endpoint. The manuscript keeps those categories visible because a useful training metric can become unsafe if it is presented as a clinical claim.

The corrected metadata errors strengthen the audit trail because they remain visible. The app export initially contained duplicate or mismatched row identifiers and build metadata. Those defects were caught against the locked row guide, repaired in the corrected Sheet, and preserved in the QC Summary and provenance note. A reviewer should be able to start with one result row and trace it back to the JSON export, video filename, app build, model version, condition assignment, correction status, and QC status.

The conflict of interest also affects methods. The same company that built the connected workflow is represented among the authors. That does not invalidate the dataset, but it raises the burden for traceability, locked files, prespecified thresholds, independent blinded review, adjudication rules, and visible failure rows. The observed package meets the corrected-package completeness part of that burden. It does not yet meet the reviewer-agreement or report-quality part.

The next validation step should test measurement validity directly. Blinded video reviewers should adjudicate first model-detected glottic target, acceptable view, view loss, no-target false detection, and report quality before reviewing generated reports. The controlled source-video record also needs an independent integrity manifest with run ID, filename, duration, byte size, checksum, storage location, access log, and reviewer verification status before the package can support measurement-validity claims. Future operator-facing studies should prespecify workload, usability, trust calibration, distraction, override and suppression behavior, trainer interpretation, and local workflow endpoints, using established workload or usability instruments when appropriate [19,20]. Cadaver testing should add realistic anatomy, contamination, lighting variation, usability burden, and safety review. Any clinical quality-improvement feasibility work should begin only after local governance, deidentification, access control, trainer oversight, and data-retention rules are defined. The value of this study is that it creates the run-level evidence chain those later studies need.

### 6.1 Limitations

This 30-run manikin corrected-package study has one overriding limitation: it is formative and simulation based. It supports measurement-readiness and traceability claims, not clinical efficacy, training effectiveness, autonomous guidance, tube-placement confirmation, or patient-outcome claims.

The 30-run manikin design also has inherent limits. The corrected export package can test export completeness, video linkage, app/model metadata, run-ID traceability, no-target behavior, exported esophageal-related checklist fields, and descriptive score outputs. It cannot by itself test blinded timing agreement, event-count agreement, report quality, trainer utility, trainee learning, workload, trust, behavior change, esophageal-intubation detection, or clinical efficacy. Performance in simulator video may differ from cadaver or clinical airway video because lighting, contamination, camera motion, anatomy, urgency, and team communication can change the visual distribution. The simulator runs did not include blood, secretions, edema, distorted anatomy, physiologic deterioration, or full team stress; the C4 degradation condition was a visual-motion stressor, not a clinical chest-compression physiology model.

The author-affiliated implementation creates a conflict that requires active management. The same company that built the connected workflow is represented among the authors. The study design reduces that risk through locked run files, prespecified thresholds, blinded video review before report review, adjudication rules, reviewer-independence disclosure, and a traceable run-level supplement. These safeguards do not remove the conflict, but they make the measurement claim auditable. The corrected Sheet shows why that matters: metadata errors in the app export were caught, repaired against the locked row guide, and preserved in the QC Summary rather than hidden.

The current public package does not include independent row-placement adjudication or an independent reviewer signature for the correction process. Row placement is author-attested. Source videos and source-video integrity records remain controlled-access artifacts rather than public repository files, and no still or video supplement is submitted with this upload. These access decisions do not change the corrected-package completeness results, but they set a hard boundary: the package is ready for blinded review, not a substitute for blinded review.

The composite score, screen-view POGO proxy, screen-view Cormack-Lehane proxy, navigation metrics, and technique bands are not validated clinical measures. They are candidate training-feedback measures and remain exploratory until tested against blinded labels, trainer judgment, workload effects, and prospective training outcomes. Their role in this manuscript is source-linked reporting, not performance certification.

## 7 Conclusion

In this corrected 30-run manikin JSON extraction, all rows contained parseable Study Metrics JSON, a companion videoFilename, run-named video-upload status, build 75 app metadata, model version 31, and unique studySessionId values aligned to the assigned condition and replicate after documented metadata reconciliation (30/30 for each completeness field; descriptive exact binomial 95% CI, 88.4% to 100.0%). The JSON target-state flag was true in target-condition rows (25/25; descriptive exact binomial 95% CI, 86.3% to 100.0%) and false in C6 no-target controls (5/5; descriptive exact binomial 95% CI, 47.8% to 100.0%) with zero glottic-detected frames, and no exported esophageal-related checklist fields were positive in the JSON exports.

These findings support a bounded formative post-reconciliation package-completeness claim: simulated video laryngoscopy attempts can be represented as specified, video-linked computer-vision procedural telemetry fields for debrief, audit, and future blinded validation after documented reconciliation. They do not establish native uncorrected export reliability, computer-vision detection accuracy, clinical efficacy, training effectiveness, autonomous guidance, tube-placement confirmation, reviewer agreement, report quality, or patient outcomes. The next evidentiary step is blinded video review and report-quality scoring against the frozen run package.

## Data Availability

No patient-level data or prospective clinical study data are analyzed in this manuscript. The manuscript reports a corrected 30-run manikin JSON/Sheet evidence package after documented metadata reconciliation.
The frozen observed-data package is publicly archived in the clean public manuscript repository as evidence-package-2026-06-20 at public release commit 5b337fe9d642ec31e3044096b56f6bca3c349869. The package directory is available at https://github.com/IntuBlade/intublade-procedural-guidance-paper-public/tree/5b337fe9d642ec31e3044096b56f6bca3c349869/evidence-package-2026-06-20. That public release commit is the cited observed-data snapshot for this submission and supersedes earlier development-package archives from the private working repository. Later manuscript-formatting commits in the same public repository do not change the frozen observed-data package unless separately stated.
Public reproducibility is at the corrected JSON, manifest, QC, correction-ledger, package-hash, package-check, and figure-generation level. The public sequence image is visual context only and is not an analyzed outcome. No public source-video supplement is released with this submission; repository placeholders, if present, are administrative transparency markers only and are not supplement media. The source-media integrity manifest is not a submitted supplement and is not presented as completed public source-media verification. Source videos, additional still images, and report screenshots remain controlled-access artifacts; controlled access can be arranged through the corresponding author where permitted. The public repository does not support independent reproduction of video-to-JSON extraction, model inference, or source-video content verification.
The clean public manuscript repository is available at https://github.com/IntuBlade/intublade-procedural-guidance-paper-public. It preserves the evidence-package source files, result-figure source, and package-check script used for this revision. The iOS implementation repository is not publicly available at this stage.

## Declarations

### Funding

No external funding was received for this manuscript. IntuBlade Co. provided the device, software, repository infrastructure, controlled storage, analysis infrastructure, and author or staff time.

### Competing interests

Andrew Napier is the founder of IntuBlade and has a financial interest in the company. Serge Klement and Ben Fedeles are affiliated with IntuBlade Co. Company-affiliated authors designed, collected, reconciled, analyzed, and drafted the study using an author-affiliated implementation. Completed conflict-management steps for this report were a locked run package, prespecified thresholds, run-level traceability, explicit correction logging for app-export metadata, source-linked evidence artifacts, and public repository archiving. Planned conflict-management steps before reporting measurement validity are blinded video labeling before report review, adjudication rules, and reviewer-independence disclosure.

### Ethics and governance

This manikin validation does not include patient data or patient-care activity. Controlled source videos remain outside the public repository under corresponding-author custody and are restricted to named study reviewers or auditors under a documented request, access log, defined retention period, and non-redistribution rule. Future manikin data collected with human operators or identifiable recordings will require appropriate local simulation-lab, institutional, privacy, and data-security review, including consent where applicable and explicit rules for whether recordings may be reused for model development. Cadaver or clinical video capture requires separate governance review.

### Data availability

The frozen observed-data package is publicly archived in the clean public manuscript repository as evidence-package-2026-06-20 at public release commit 5b337fe. That public release commit is the cited observed-data snapshot for this submission and supersedes earlier development-package archives from the private working repository. Later manuscript-formatting commits in the same public repository do not change the frozen observed-data package unless separately stated. The package includes the corrected 30-run Sheet export as CSV and XLSX, the run-level supplement, 30 corrected Study Metrics JSON exports, the run-named video manifest, QC Summary, structured correction ledger, package hash manifest, condition protocol, data dictionary, provenance note, package telemetry transparency card, governance-readiness note, observed-results summary, package-check script, source-media decision notes, and one representative four-panel simulator-airway guidance sequence for visual context only. The run-named source videos are stored in the study Drive capture folder for this work and are represented in `run_manifest.csv`; the GitHub evidence package stores filenames, corrected JSON exports, manifest linkage, QC records, package checks, and the representative visual-context sequence image rather than source-video bytes. Public reproducibility is at the corrected JSON, manifest, QC, correction-ledger, package-hash, package-check, and figure-generation level. At this revision, no still-image or video supplement is submitted as an analyzed outcome. The source-media integrity manifest is an administrative tracker and is not presented as completed public source-media checksum verification. Full source-video review, public source-video release, or independent reproduction of video-to-JSON extraction would require a separately completed media-integrity manifest, access or release decision, and any required governance review.

### Code availability

The clean public manuscript and evidence-package repository is the code and package repository for this paper. It includes the corrected evidence package, generated figure outputs, figure metadata, run-level supplement, result-figure source (`build_observed_results_figures_2026_06_20.py`), and package-check script (`check_observed_package.py`). The production iOS app implementation repository that generated the study exports is separate from this manuscript repository and is not released here; therefore, the public package supports audit of the exported telemetry package but not independent re-execution of app inference. The reproducibility record for the validation build is app build 3.3.0 (75), source revision b94cd63, model version 31, detection threshold 0.1, and the archived telemetry schema.

### Author contributions

Andrew Napier contributed the clinical concept, protocol framing, implementation context, interpretation limits, and manuscript drafting. Serge Klement contributed implementation context, software architecture, and technical review. Ben Fedeles contributed clinical review, safety-boundary review, and manuscript revision.

## Acknowledgments

No acknowledgments.

## AI assistance

AI-assisted writing and coding tools were used to support manuscript editing, figure-generation code review, formatting checks, and revision planning. The authors reviewed, revised, and approved the manuscript, analysis code, figures, evidence-package language, and all scientific claims.

## Notes

### Clinical Protocols

https://github.com/IntuBlade/intublade-procedural-guidance-paper-public/blob/5b337fe9d642ec31e3044096b56f6bca3c349869/evidence-package-2026-06-20/protocol_setup.md

### Author Declarations

This work used only manikin/simulator data generated for this project, corrected JSON/Sheet exports, and public repository files. No patient-level data or prospective clinical study data were analyzed. The public package provides corrected JSON exports, manifests, QC records, package hashes, scripts, protocol files, and a representative visual-context sequence. Source videos remain controlled-access under corresponding-author custody and are represented publicly by filename and manifest linkage rather than by public video bytes.

